# An evidence-based data science perspective on the prediction of heart failure readmissions

**DOI:** 10.1101/2021.05.10.21256926

**Authors:** Kenneth J. Locey, Thomas A. Webb, Bala Hota

## Abstract

The prevention of unplanned 30-day readmissions of patients discharged with a diagnosis of heart failure (HF) remains a profound challenge among hospital enterprises. Despite the many models and indices developed to predict which HF patients will readmit for any unplanned cause within 30 days, predictive success has been meager. Using simulations of HF readmission models and the diagnostics most often used to evaluate them (C-statistics, ROC curves), we demonstrate common factors that have contributed to the lack of predictive success among studies. We reveal a greater need for precision and alternative metrics such as partial C-statistics and precision-recall curves and demonstrate via simulations how those tools can be used to better gauge predictive success. We suggest how studies can improve their applicability to hospitals and call for a greater understanding of the uncertainty underlying 30-day all-cause HF readmission. Finally, using insights from sampling theory, we suggest a novel uncertainty-based perspective for predicting readmissions and non-readmissions.

---

Over the past decade, hospitals have been motivated to decrease 30-day all-cause readmissions among heart failure (HF) patients.^1^ These HF readmissions (HFR) are often deemed preventable and failing to decrease them carries financial penalties.^1^ Consequently, to aid hospital enterprises in reducing HFR, hundreds of studies have used or developed indices and models to predict HFR, 30-day all-cause or otherwise.^2-4^ These HFR studies have varied greatly in their readmission time frames (30d – 1yr) and methodologies (risk indices, statistical models, machine learning), as well as in the data underpinning their predictions (clinical, administrative, psychosocial).^4-8^ However, the degrees of performance and applicability needed to make transformative progress in predicting HFR have been notoriously difficult to achieve ^4,9^, begging the question of why.

While the variation in methods, data, and performance among HFR studies has been thoroughly reviewed by others ^3,4,10^, few explanations have been given for the shared lack of success or the commonalities that have contributed to it. In the current work, we identify and discuss common factors contributing to the difficulty in predicting HFR, 30-day all-cause or otherwise. We demonstrate how the choice of diagnostic metrics, data, and models can not only impede progress but also prevent the development of useful tools and actionable solutions. In each case, we suggest alternatives and potential paths forward. Finally, we recast the prediction of HFR with an uncertainty-based perspective.

## ROC curves and C-statistics

HFR studies often demonstrate their success via receiver operating characteristic (ROC) curves.^4,7,10,11^ ROC curves relate the true positive rate (TPR) to the false positive rate (FPR) as a classifier is applied across diagnostic thresholds (Table 1, Fig 1a). In HFR studies, thresholds are often measures of risk or probabilities above which an index discharge is classified as resulting in readmission. ROC curves represent the trade-off between correctly classifying discharges that result in readmission and incorrectly classifying discharges that do not. To quantify their ROC results, HFR studies use C-statistics (aka C-index) representing the area under the ROC curve (AUROC).^4,7,10,11^ These C-statistics range from 0 to 1, with values near 0.5 being no better than random and values greater than 0.9 signifying outstanding diagnostic power (Fig 1a).^12^ For context, C-statistics (i.e., AUROC) for 30-day all-cause HFR studies typically range between 0.5 and 0.7.^7,10,11,13-18^

**Table 1.** Diagnostic measures related to receiver operating characteristic (ROC) curves and precision-recall curves (PRC). P = positives = readmissions. N = negatives = non-readmissions. TP = True positives, FP = False positives, TN = True negatives, FN = False negatives.

| Measure | Equation | Description relative to HF readmissions (HFR) | Note |
| --- | --- | --- | --- |
| True positive rate (TPR), sensitivity, recall | $TPR = TP/P = TP/(TP+FN)$ | Fraction of positives correctly classified | No established minimum acceptable TPR for HFR |
| False positive rate (FPR), False alarm rate | $FPR = FP/N = FP/(FP+TN)$ | Fraction of non-readmissions incorrectly classified as positives | No established maximum acceptable FPR for HFR |
| True negative rate (TNR), specificity | $TNR = TN/N = TN/(TN + FP)$ | Fraction of non-readmissions correctly classified as negatives | Rarely used in HFR studies |
| False negative rate (FNR) | $FNR = FN/P = FN/(FN+TP)$ | Fraction of readmissions incorrectly classified as negatives | Rarely used in HFR studies |
| Precision; positive predictive value (PPV) | $PPV = TP/(TP+FP)$ | Fraction of predictions for readmission that were correct. | Rarely used in HFR studies |
| Negative predictive value (NPV) | $NPV = TN/(TN+FN)$ | Fraction of predictions for non-readmission that were correct. | Rarely used in HFR studies |

**Figure 1.**
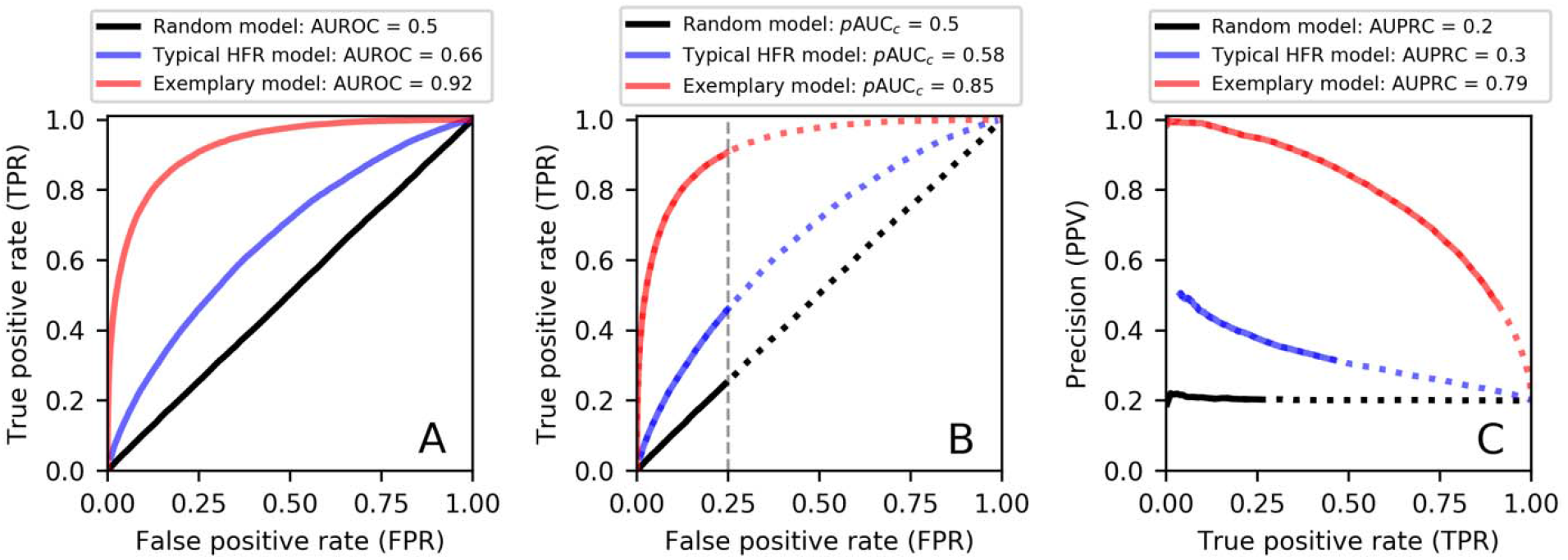
Receiver operating characteristic (ROC) curves and precision-recall curves (PRC) resulting from analyzing a single simulated dataset of 10^5^ binary outcomes (20% positives, 80% negatives) with three hypothetical models. These include a model programmed to make unbiased random guesses (black lines), a model programmed to produce results typical of many HFR models (blue lines), and a model programmed to represent an HFR model of exceptionally high diagnostic power (red lines). See Appendix 1 for modeling details. **A)** Examples of ROC curves and AUROC values. As expected, the random model approximates the 1:1 line (AUROC = 0.5). **B)** Same ROC curves as in A but showing partial AUROC (*p*AUC_*c*_) values using a maximally acceptable false positive rate of 0.25. Solid lines represent the range of FPR below 0.25. Note how *p*AUC_*c*_ = AUROC = 0.5 for the random model, but *p*AUC_*c*_ < AUROC for other models. **C)** PRCs and their AUPRC values. Solid lines represent FPR values below 0.25, as in B. This figure can be recreated using the supplemental Fig1.py python script.

ROC curves and C-statistics provide a standard way to report results and compare studies. However, these metrics provide an incomplete view of diagnostic success and can hide diagnostic failures. For example, only a small portion of the ROC curve may be useful because only a small range of FPR may be acceptable.^19^ Among HFR studies, FPR often exceeds 0.25 before the TPR exceeds 0.5.^11,20,21^ Simply put; the false alarm rate often reaches 25% before 50% of discharges resulting in readmission are correctly classified. If an FPR greater than 0.25 were unacceptable, then a C-statistic or AUROC based on the full ROC curve would be misleading. Consequently, HFR studies may benefit from partial AUROC, i.e., the area under the ROC curve corresponding to an acceptable range of FPR (Fig 1b). Partial AUROC measures and corresponding partial C-statistics have been developed by others, included into analytical libraries, and have been used in other healthcare studies.^19,22^

Whether using partial C-statistics for a maximally acceptable FPR or not, there is a more deeply concerning detriment to using ROC curves as a standard of performance. Specifically, ROC curves do not reveal the probability of making a correct positive prediction (i.e., precision) and hence, whether a prediction for readmission can be trusted. Likewise, the degree of diagnostic failure obscured by ROC curves can grow as data become imbalanced, i.e., as non-readmissions outnumber readmissions. We address these issues below.

## Precision and imbalanced data

Also known as the positive predictive value (PPV), precision is the fraction of positive predictions that were actually correct (Table 1). Surprisingly, many HFR studies never mention or measure precision.^2-4,7,8,13-16,18,20,23-27^ However, without precision there is little way to establish confidence in whether or not a prediction for readmission can be trusted. Among the comparatively few studies that report it, precision typically ranges from 0.09 to 0.44, meaning that 56 – 91% of predictions for readmission are often incorrect.^3,21,24,28,29^ If hospitals acted on the results of such predictions, they would invest their limited time and resources into expectations that are, more often than not, false alarms.

Precision and TPR are often inversely related; an increase in one will produce a decrease in the other.^19^ Consequently, if HFR studies give greater attention to precision, they will inevitably find that models with impressive ROC curves can be highly imprecise. Simulations can reveal how this occurs (Fig 1a-c, Appendix 1). In the analyses behind Figure 1, applying a hypothetical high performing model to simulated HFR outcomes produced an AUROC of 0.92 and a TPR of 0.9 at a maximally allowed FPR of 0.25 (Fig 1b). If this were a real model applied to real HFR data, the result would be exemplary among HFR studies. Yet, the corresponding precision of this model was only 47% (Fig 1c). Another hypothetical model designed to be similar in performance to real HFR models (AUROC = 0.66) produced a TPR of 0.47 at an FPR of 0.25, with a precision of only 32% (Fig 1a-c).

The tradeoff between accurately capturing readmissions and making reliable predictions of readmission (TPR vs. precision) is one complicating factor for predictive HFR studies. The other factor is class imbalance, i.e., large differences in the numbers of readmissions and non-readmissions. In HFR studies, 30-day all-cause readmissions typically comprise less than 25% of HF index discharges.^2,7,13,17,18,23^ This class imbalance poses challenges for machine learning algorithms and some HFR studies have taken measures to correct for it.^3,17^ However, class imbalance also has a less frequently acknowledged effect. Specifically, if non-readmissions greatly outnumber readmissions, then ROC curves will fail to reflect how greatly false positives outnumber true positives, causing precision to suffer in a way that ROC curves and their associated metrics cannot capture (Fig 2).

**Figure 2.**
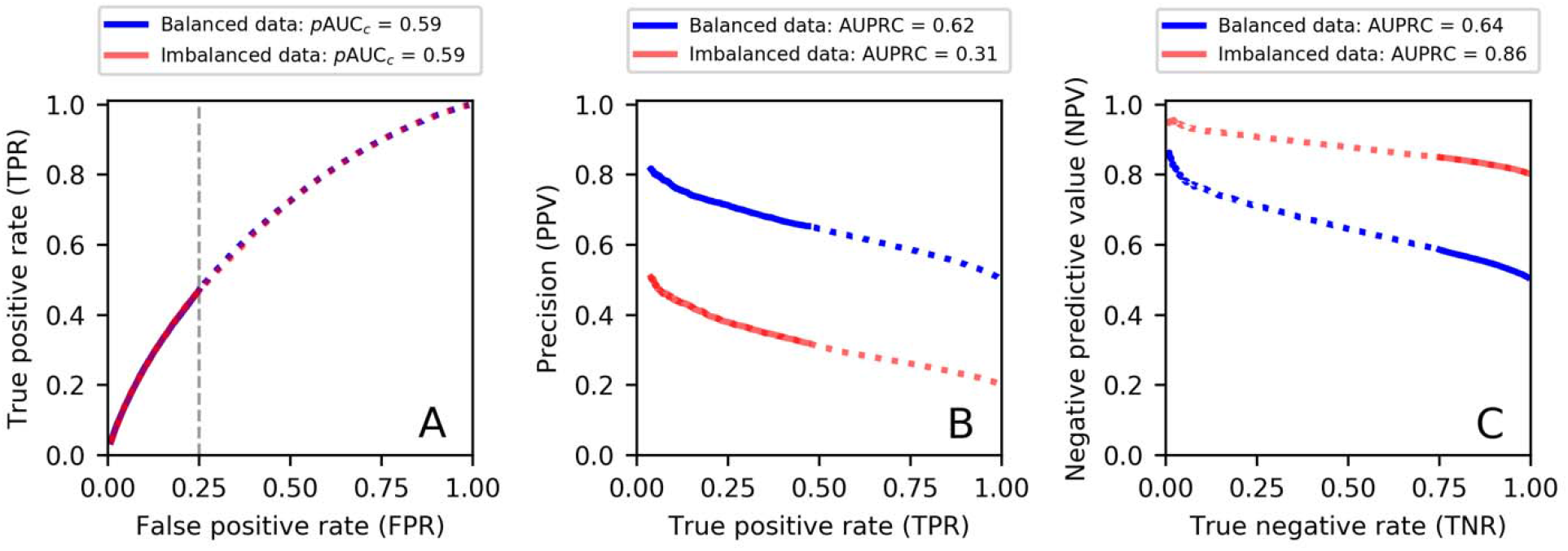
Receiver operating characteristic (ROC) curves and precision-recall curves (PRC) resulting from the application of a single model (typical of results for HFR models) to two simulated datasets (*n* = 10^5^ binary outcomes). The ratio of negative to positive outcomes was 1:1 for the balanced data set and 4:1 for the imbalanced data set. The latter ratio represents that ∼20% of index discharges in HFR datasets result in 30-day all-cause readmission. See Appendix 1 for modeling details. **A)** ROC curves typical of HFR model results; these are identical for imbalanced and balanced data sets. **B)** PRCs reveal the influence of imbalanced data on precision (i.e., positive predictive value, PPV). The AUPRC for balanced data is more than twice that for imbalanced data. **C)** PRC curves focused on negative outcomes, i.e., negative predictive value (NPV) vs. true negative rate (TNR). AUPRC for imbalanced data is substantially greater than that for balanced data. This figure can be recreated using the supplemental Fig2.py python script.

In addition to giving greater attention to precision and class imbalance, HFR studies may benefit from incorporating precision-recall curves (PRCs) (Fig 1c, Fig 2a-b). PRCs reveal the relationship between precision and TPR, thereby capturing the tradeoff between correctly classifying discharges that result in readmission and avoiding false positives. PRCs are recommended over ROC curves for use with imbalanced data and can be summarized by calculating the area under the curve (AUPRC) (Fig 1c).^19,30,31^ Noting this, a small number of HFR studies have used PRCs and reported their respective AUC values.^30,31^ As an example of the insight gained by using PRCs, we applied a hypothetical model to two data sets of simulated HFR outcomes (Fig 2a-b, Appendix 1). These two data sets only varied in their ratios of non-readmissions to readmissions (1:1, 4:1). While the resulting ROC curves were identical, PRCs reveal disparate degrees of performance.

HFR studies can provide greater insight and pursue greater rigor when developing, evaluating, and comparing predictive models by 1) using PRC’s in combination with both ROCs and partial AUROC and partial C-statistics, 2) examining a greater diversity of diagnostic measures, and by 3) establishing standards for a minimum TPR (e.g., 0.5) and a maximum FPR (e.g., 0.25). In doing so, HFR studies could attempt to optimize precision and TPR, while not exceeding an acceptable FPR.

## Meeting the needs of healthcare providers

Ideally, predictive HFR studies would be guided by the abilities of providers to readily apply an approach within time sensitive needs and without excluding patients. However, HFR studies are often vulnerable to biases, include biases in model methodology and validation, data attrition and patient exclusion, reliability in new populations, ease of implementation in clinical settings and patient management.^4,32^ Predictive HFR studies are also often retrospective, i.e., using data that are only available after the point of intervention has passed.^16,32^ Consequently, without considering the needs and limitations of healthcare providers, HFR studies can fail to foresee biases that impede the ability of hospitals to implement an approach. In particular, the advancing sophistication of predictive HFR studies may exacerbate these biases.

Predictive HFR studies have begun using advanced forms of deep learning such as convolutional neural networks, deep-unified neural networks, and hybrid topic recurrent neural networks.^31-34^ These tools represent some of the most powerful algorithms that data science has to offer but require high levels of technical expertise in the design of network architecture, choice of hyperparameters, and model validation. When married to massive amounts of patient data, advanced forms of machine learning may require computational resources that most hospitals lack. And, when requiring data that many hospitals may not readily have (e.g., complete assays of laboratory results and clinician notes), advanced machine learning methods can be self-limiting in regards to practical application.^32^

Authors of HFR studies could increase their impact by aiming for real-time predictions and by avoiding the above biases. ^4,16,32^ Going further, studies could increase their usefulness to hospitals by collaborating with clinical teams to clarify how an approach can be used in practice, if at all, and at what point in patient management. Likewise, predictive HFR studies could increase their usefulness among researchers and hospital data scientists by adopting higher standards of reproducibility. In particular, HFR studies rarely make their analytical source code and permissible data available via public repositories (e.g., via GitHub). By offering public repositories containing permissible data (e.g., simulated pseudo-data) and well-documented source code built with open-source tools (e.g., Python, R), HFR studies can enable others to use, test, and improve upon existing methods.

## Inherent uncertainty of 30-day all cause HFR

Predicting HF-related mortality, HFR beyond 30 days, or predicting which patients have HF often leads to greater success than predicting 30-day all-cause HFR.^4,5,10,13,14,20,21,31,33,35^ As opposed to using a strong diagnostic such as elevated cardiac troponin to predict whether a patient has HF or will expire within one year, predicting 30-day all-cause HFR often means using data with a weak signal to predict whether a patient will readmit for any unplanned cause within a relatively short period of time. The inherent uncertainties of this problem make it a grand predictive challenge and whether it outstrips the classification problems to which machine learning is often successfully applied is unknown. However, the most sophisticated deep learning algorithm will fail when making predictions about random outcomes (e.g., coin flips). Though prior studies have shown that 30-day all-cause HFR is not entirely random, the overall failure to breach C-statistics above 0.8 and precision above 0.5, despite greatly varied data and increasingly advanced methods, suggests a degree of stochasticity that is poorly understood and rarely discussed.

Authors have suggested that increasingly sophisticated machine learning applied to a greater volume of relevant data will improve 30-day all-cause HFR predictions.^9,11,29^ Given the march of advancing methods and increasingly large data sets, many have invested in this assumption. But, to our knowledge, that assumption has never been tested and it remains to be seen what data are most useful for predicting 30-day all-cause HFR and what models will push the field beyond incremental success. It seems clear, however, that hospitals are penalized on what seems to be the most difficult aspect of HFR to predict. Consequently, progress towards predictive power and the continuation of policies that penalize against readmissions need a clearer understanding of the inherent stochasticity of 30-day all-cause HFR.

## Missed opportunities

The common goal of HFR studies is to predict which HF patients will readmit or, at least, which are at greatest risk. Considering the precision and accuracy needed and the urgent needs of hospitals, additional goals should be considered. For example, HF patients that do not readmit may be easier to predict than those that do. Among HFR studies that report positive predictive values (PPV; precision) and negative predictive values (NPV), only 19% – 44% of predictions for readmission were correct, while 80 – 96% of predictions for non-readmission were correct.^21,24,28^ However, no HFR study has explicitly asked whether or why predicting negative outcomes (non-readmissions) is easier than predicting positive ones (readmissions) or how the ability to predict non-readmissions can be used.

Statistical sampling theory and the inspection of diagnostic metrics (TPR, FPR, PPV, NPV) suggests that the class imbalance of HFR data should cause false negatives to have less of an impact on the probability of predicting a non-readmission (NPV) than false positives have on PPV (Fig 2b-c, Table 1). In short, confident predictions for non-readmissions may be naturally easier to achieve. Additionally, and also related to class imbalance, a given HFR data set is bound to be richer in information on negative outcomes (non-readmission) than positive ones, because non-readmissions are more common than readmissions. Taken together, predicting non-readmissions should be less sensitive to misclassification and offer more data on which confident predictions can be based.

Given that success towards predicting HFR has advanced little in the past decade with respect to the sophistication of the tools applied or the data used, a more strategic use of predictive analytics is needed. To this end, we suggest an uncertainty-based perspective. Specifically, if predictions for non-readmission are often precise (e.g., NPV > 0.8), then predictions with a high probability of non-readmission should aptly be treated as “high certainty for non-readmission”. Likewise, because predictions for readmission are typically incorrect (i.e., PPV < 0.4), predictions normally regarded as indicating “high risk” should more aptly be treated as predictions of “high uncertainty”.

Stratifying patients by uncertainty rather than presumably greater risk may allow hospitals to more effectively distribute limited resources (e.g., outpatient care, phone calls, virtual visits). Rather than overstretch resources by assuming that patients of presumably high risk are truly at high risk, hospitals could look deeper into their “high uncertainty” cohort for signs of greater certainty. Additionally, perhaps filtering out the “high certainty for non-readmission” cohort from HF discharge data will help correct the class imbalance in HF data sets and allow algorithms to focus their training on patients of greater uncertainty. Integrating precise predictions for non-target cases is bound to be more useful than ignoring them and, when “high risk” patients readmit at a frequency less than 50%, it is almost certainly more appropriate and useful to consider them as patients of high uncertainty.

## Beyond HFR

The challenges of predicting 30-day all-cause readmissions and the penalties imposed on hospitals for not decreasing them, extend beyond patients with an HF discharge diagnosis. Hospitals face the same challenges for patients discharged with pneumonia, acute myocardial infarction, and other clinical conditions or procedures.^1^ We suspect that predicting 30-day all-cause readmissions for most any discharge diagnosis is likely fraught with similar challenges. In each case, hospitals need easily integrated tools that make real-time predictions for strongly stochastic events from all-to-often weakly diagnostic data, and quickly enough to make informed decisions.

Beyond aiming for larger datasets and increasingly sophisticated models, we expect studies that eventually overcome these predictive challenges will be aided by: 1) a greater focus on precision (PPV, NPV), 2) a better choice of diagnostic metrics (e.g., PRCs, partial AUCs), 3) greater attention to practical application and reproducibility, 4) an exploration of additional goals (e.g., prediction of non-readmissions) and perhaps, 5) the adoption of an uncertainty-based perspective that clarifies the inherent stochasticity of 30-day all-cause readmissions and, likewise, that allows researchers and hospitals to best deal with it.

## Supporting information

Supplemental python script to generate figure 1

Supplemental python script to generate figure 2

## Data Availability

All analyses in our work were performed via computer simulations. No clinical or other data is associated.

