## Appendix 1 for "An evidence-based data science perspective on the prediction of heart failure readmissions"

**Appendix 1: Descriptions of modeling used in Figures 1 and 2 of the main manuscript**

**Models underpinning figure 1.**

We used simulation models based on random draws from probability distributions to construct hypothetical receiver operating characteristic (ROC) curves and hypothetical precision-recall curves (PRCs). We first generated a simulated data set of 10^5^ binary outcomes (20% positives, 80% negatives), resulting in a list of 0.2 • 10^5^ ones and 0.2 • 10^5^ zeros, with ones representing hypothetical readmissions and zeros representing hypothetical non-readmissions. Here, we denote this set as *S*. We then simulated the classification of outcomes according to three hypothetical models, each of which produced hypothetical probabilities of readmission ranging between 0 and 1.

Model 1: Unbiased random guesses – This model classifies each outcome in *S* using random draws from a uniform distribution. Analysis of the resulting outcomes via receiver operating characteristic curves would produce a C-statistic of 0.5 and demonstrate the expected area under the precision-recall curve, i.e., 0.2.

Model 2: Typical HFR model – This model was based on random draws from a Gaussian distribution. For each outcome in S, the classification was obtained by taking a single random draw from a Gaussian distribution with a mean equal to the value of the outcome (0 or 1), and a standard deviation equal to 1.7. This sampling procedure was more likely than an unbiased random guess (e.g., Model 1) to make a correct prediction but because of the relatively high standard deviation, produced results similar to those often observed for predictive heart failure readmission models.

Model 3: Exceptionally accurate HFR model – As with Model 2, this model was based on random draws from a Gaussian distribution. For each outcome in S, the classification was obtained by taking a single random draw from a Gaussian distribution with a mean equal to the value of the outcome (0 or 1), and a standard deviation equal to 0.5. This sampling procedure was more likely than Model 1 and Model 2 to make correct predictions because of its relatively low standard deviation. Consequently, it produced hypothetical results that were exceptionally accurate as compared to nearly all predictive heart failure readmission models.

For each model, the observed outcomes (i.e., *S*) were compared to the predicted outcomes (hypothetical probabilities of readmission) using typical ROC and PRC analyses. Specifically, we applied thresholds of increasing value (ranging between 0 and 1) to the list of hypothetical probabilities, with each prediction either indicating a readmission (1) or a non-readmission (0) depending on whether the hypothetical probability was equal to or greater than the threshold value. These models and subsequent analysis are included in the supplemental **Fig1.py** python script.

**Models underpinning figure 2.**

We used a simulation model based on random draws from a Gaussian distribution to construct hypothetical receiver operating characteristic (ROC) curves and hypothetical precision-recall curves (PRCs). We first generated two simulated data sets of 10^5^ binary outcomes. The first data set was characterized by 20% positives and 80% negatives, i.e., a list containing 0.2 • 10^5^ ones and 0.2 • 10^5^ zeros with ones representing hypothetical readmissions and zeros representing hypothetical non-readmissions. The second data set was comprised of 50% positives and 50% negatives, i.e., a list of 0.5 • 10^5^ ones and 0.5 • 10^5^ zeros. We denote this set as *S*.

For each outcome in *S*, the hypothetical classification was obtained by taking a single random draw from a Gaussian distribution with a mean equal to the value of the outcome (0 or 1), and a standard deviation equal to 1.7. This sampling procedure was more likely than an unbiased random guess to make a correct prediction but because of the relatively high standard deviation, produced results similar to those often observed for predictive heart failure readmission models. The observed outcomes (i.e., *S*) were compared to the predicted outcomes (hypothetical probabilities of readmission) using typical ROC and PRC analyses. Specifically, we applied thresholds of increasing value (ranging between 0 and 1) to the list of hypothetical probabilities, with each prediction either indicating a readmission (1) or a non-readmission (0) depending on whether the hypothetical probability was equal to or greater than the threshold value. These analyses are included in the supplemental **Fig2.py** python script.
