## Supplemental python script to generate figure 2 for "An evidence-based data science perspective on the prediction of heart failure readmissions": Fig2.py

```
import pandas as pd
import numpy as np
import sys
from scipy import stats
import os
import warnings
import random
from sklearn.metrics import auc
from sklearn.metrics import roc_auc_score
import matplotlib.pyplot as plt

mydir = os.path.expanduser("/Volumes/RushProjects/AvoidableReadmits")

import pAUCc as PAUCC # See: https://github.com/Big-Life-Lab/partial-AUC-C/tree/main/Python3.7

Ns = [0] * 80000
Ps = [1] * 20000
obs_imb = Ns + Ps

Ns = [0] * 50000
Ps = [1] * 50000
obs_bal = Ns + Ps


def get_pred(pop, s):
    pred = []
    s1 = s
    for n in pop:
        x = np.random.normal(loc=n, scale=s1)
        pred.append(x)
        
    return pred


def get_lists(obs, exp):
    tpr_ls = []
    tnr_ls = []
    fpr_ls = []
    fnr_ls = []
    ppv_ls = []
    npv_ls = []
    ct_ls = []
    tp_ls = []
    fp_ls = []
    tn_ls = []
    fn_ls = []

    cut_ls = np.linspace(4.0, -4.0, 3000).tolist()

    for c in cut_ls:
        
        tp, fp, tn, fn = 0,0,0,0
        tpr, fpr, fnr, tnr = 0,0,0,0
        ppv, npv = 0, 0

        for i, o in enumerate(obs):
                
            e = exp[i]
            
            if o == 0 and e < c: 
                tn += 1
            elif o == 0 and e >= c:
                fp += 1
            elif o == 1 and e >= c:
                tp += 1
            elif o == 1 and e < c:
                fn += 1
            else:
                print('something is not adding up')
                sys.exit()
            
        try: tpr = tp/(tp + fn)
        except: tpr = np.nan
        try: fpr = fp/(fp + tn)
        except: fpr = np.nan
        try: tnr = tn/(tn + fp)
        except: tnr = np.nan
        try: ppv = tp/(tp + fp)
        except: ppv = np.nan
        try: npv = tn/(tn + fn)
        except: npv = np.nan
        try: fnr = fn/(fn + tp)
        except: fnr = np.nan
        
        tpr_ls.append(tpr)
        fpr_ls.append(fpr)
        tnr_ls.append(tnr)
        fnr_ls.append(fnr)
        ppv_ls.append(ppv)
        npv_ls.append(npv)
        ct_ls.append(c)
        
        tp_ls.append(tp)
        fp_ls.append(fp)
        tn_ls.append(tn)
        fn_ls.append(fn)

        
    return tpr_ls, tnr_ls, fpr_ls, fnr_ls, ppv_ls, npv_ls, ct_ls, tp_ls, fp_ls, tn_ls, fn_ls


bal_pred = get_pred(obs_bal, 1.7)
imb_pred = get_pred(obs_imb, 1.7)

results_bal = get_lists(obs_bal, bal_pred)
results_imb = get_lists(obs_imb, imb_pred)


lw, rows, cols, a = 2, 2, 2, 0.6
fig = plt.figure(figsize=(8, 8))

fs = 8
#bbta = (-0.15, 1.02, 1.25, 0.3)
bbta = (-0.05, 1.02, 1.1, 0.3)

#### a. ROC curves and partial AUC
ax = fig.add_subplot(3, 3, 1)

plt.vlines(0.25, 0, 1.1, alpha=0.5, color='0.2', linestyles='dashed', linewidth=lw-1)

tpr_ls, tnr_ls, fpr_ls, fnr_ls, ppv_ls, npv_ls, ct_ls, tp_ls, fp_ls, tn_ls, fn_ls = results_bal
tpr, fpr = [], []
tpr_part, fpr_part = [], []
for i, val1 in enumerate(tpr_ls):
    val2 = fpr_ls[i]
    if val1 >= 0 and val1 <= 1.0 and val2 >= 0 and val2 <= 0.25:
        tpr_part.append(val1)
        fpr_part.append(val2)
    if val1 >= 0 and val1 <= 1.0:
        tpr.append(val1)
        fpr.append(val2)
pAUCc = PAUCC.concordant_partial_AUC(fpr_part, tpr_part)
pAUCc = pAUCc[3]
plt.plot(fpr_part, tpr_part, c = 'b', label='Balanced data: ' + r'$p$' +'AUC'+r'$_{c}$' + ' = ' +str(np.round(pAUCc, 2)), linewidth=lw)
plt.plot(fpr, tpr, c = 'b', linewidth=lw, linestyle=':')

tpr_ls, tnr_ls, fpr_ls, fnr_ls, ppv_ls, npv_ls, ct_ls, tp_ls, fp_ls, tn_ls, fn_ls = results_imb
tpr, fpr = [], []
tpr_part, fpr_part = [], []
for i, val1 in enumerate(tpr_ls):
    val2 = fpr_ls[i]
    if val1 >= 0 and val1 <= 1.0 and val2 >= 0 and val2 <= 0.25:
        tpr_part.append(val1)
        fpr_part.append(val2)
    if val1 >= 0 and val1 <= 1.0:
        tpr.append(val1)
        fpr.append(val2)
pAUCc = PAUCC.concordant_partial_AUC(fpr_part, tpr_part)
pAUCc = pAUCc[3]
plt.plot(fpr_part, tpr_part, c = 'r', label='Imbalanced data: ' + r'$p$' +'AUC'+r'$_{c}$' + ' = ' +str(np.round(pAUCc, 2)), alpha = a, linewidth=lw)
plt.plot(fpr, tpr, c = 'r', alpha = a, linewidth=lw, linestyle=':')

plt.legend(bbox_to_anchor=bbta, loc=10, ncol=1, mode="expand",prop={'size':fs-2})
plt.ylim(0, 1.01)
plt.xlim(0, 1.0)
plt.xlabel('False positive rate (FPR)', fontsize=fs)
plt.ylabel('True positive rate (TPR)', fontsize=fs)
ax.tick_params(axis='both', which='major', labelsize=fs)
plt.text(0.85, 0.05, 'A', fontsize=fs+5)        
        
        
        
        
#### b. PRCs corresponding to ROCs in a
ax = fig.add_subplot(3, 3, 2)

tpr_ls, tnr_ls, fpr_ls, fnr_ls, ppv_ls, npv_ls, ct_ls, tp_ls, fp_ls, tn_ls, fn_ls = results_bal
tpr, ppv = [], []
tpr_part, ppv_part = [], []
for i, val1 in enumerate(tpr_ls):
    val2 = ppv_ls[i]
    val3 = fpr_ls[i]
    if val1 >= 0 and val1 <= 1.0 and val2 >= 0 and val2 <= 1.0 and val3 >= 0 and val3 <= 0.25:
        tpr_part.append(val1)
        ppv_part.append(val2)
    if val1 >= 0 and val1 <= 1.0 and val2 >= 0 and val2 <= 1.0:
        tpr.append(val1)
        ppv.append(val2)
AUC = auc(tpr, ppv)
plt.plot(tpr_part, ppv_part, c = 'b', label='Balanced data: AUPRC = '+str(np.round(AUC, 2)), linewidth=lw)
plt.plot(tpr, ppv, c = 'b', linewidth=lw, linestyle=':')

tpr_ls, tnr_ls, fpr_ls, fnr_ls, ppv_ls, npv_ls, ct_ls, tp_ls, fp_ls, tn_ls, fn_ls = results_imb
tpr, ppv = [], []
tpr_part, ppv_part = [], []
for i, val1 in enumerate(tpr_ls):
    val2 = ppv_ls[i]
    val3 = fpr_ls[i]
    if val1 >= 0 and val1 <= 1.0 and val2 >= 0 and val2 <= 1.0 and val3 >= 0 and val3 <= 0.25:
        tpr_part.append(val1)
        ppv_part.append(val2)
    if val1 >= 0 and val1 <= 1.0 and val2 >= 0 and val2 <= 1.0:
        tpr.append(val1)
        ppv.append(val2)
AUC = auc(tpr, ppv)
plt.plot(tpr_part, ppv_part, c = 'r', label='Imbalanced data: AUPRC = '+str(np.round(AUC, 2)), alpha = a, linewidth=lw)
plt.plot(tpr, ppv, c = 'r', alpha = a, linewidth=lw, linestyle=':')

plt.legend(bbox_to_anchor=bbta, loc=10, ncol=1, mode="expand",prop={'size':fs-2})
plt.ylim(0, 1.01)
plt.xlim(0, 1.0)
plt.xlabel('True positive rate (TPR)', fontsize=fs)
plt.ylabel('Precision (PPV)', fontsize=fs)
ax.tick_params(axis='both', which='major', labelsize=fs)
plt.text(0.85, 0.05, 'B', fontsize=fs+5)


#### b. PRCs corresponding to ROCs in a
ax = fig.add_subplot(3, 3, 3)

tpr_ls, tnr_ls, fpr_ls, fnr_ls, ppv_ls, npv_ls, ct_ls, tp_ls, fp_ls, tn_ls, fn_ls = results_bal
tnr, npv = [], []
tnr_part, npv_part = [], []
for i, val1 in enumerate(tnr_ls):
    val2 = npv_ls[i]
    val3 = fpr_ls[i]
    if val1 >= 0 and val1 <= 1.0 and val2 >= 0 and val2 <= 1.0 and val3 >= 0 and val3 <= 0.25:
        tnr_part.append(val1)
        npv_part.append(val2)
    if val1 >= 0 and val1 <= 1.0 and val2 >= 0 and val2 <= 1.0:
        tnr.append(val1)
        npv.append(val2)
AUC = auc(tnr, npv)
plt.plot(tnr_part, npv_part, c = 'b', label='Balanced data: AUPRC = '+str(np.round(AUC, 2)), linewidth=lw)
plt.plot(tnr, npv, c = 'b', linewidth=lw, linestyle=':')

tpr_ls, tnr_ls, fpr_ls, fnr_ls, ppv_ls, npv_ls, ct_ls, tp_ls, fp_ls, tn_ls, fn_ls = results_imb
tnr, npv = [], []
tnr_part, npv_part = [], []
for i, val1 in enumerate(tnr_ls):
    val2 = npv_ls[i]
    val3 = fpr_ls[i]
    if val1 >= 0 and val1 <= 1.0 and val2 >= 0 and val2 <= 1.0 and val3 >= 0 and val3 <= 0.25:
        tnr_part.append(val1)
        npv_part.append(val2)
    if val1 >= 0 and val1 <= 1.0 and val2 >= 0 and val2 <= 1.0:
        tnr.append(val1)
        npv.append(val2)
AUC = auc(tnr, npv)
plt.plot(tnr_part, npv_part, c = 'r', label='Imbalanced data: AUPRC = '+str(np.round(AUC, 2)), alpha = a, linewidth=lw)
plt.plot(tnr, npv, c = 'r', alpha = a, linewidth=lw, linestyle=':')

plt.legend(bbox_to_anchor=bbta, loc=10, ncol=1, mode="expand",prop={'size':fs-2})
plt.ylim(0, 1.01)
plt.xlim(0, 1.0)
plt.xlabel('True negative rate (TNR)', fontsize=fs)
plt.ylabel('Negative predictive value (NPV)', fontsize=fs)
ax.tick_params(axis='both', which='major', labelsize=fs)
plt.text(0.85, 0.05, 'C', fontsize=fs+5)


plt.subplots_adjust(wspace=0.45, hspace=0.45)
plt.savefig(mydir+'/figures/Fig2.png', dpi=400, bbox_inches = "tight")
plt.close()
```
